# Associations between total MRI-visible small vessel disease burden and domain-specific cognitive abilities in a community-dwelling older-age cohort

**DOI:** 10.1101/2021.02.02.21250986

**Authors:** OKL Hamilton, SR Cox, L Ballerini, ME Bastin, J Corley, AJ Gow, S Muñoz Maniega, P Redmond, Valdés-Hernández M del C, JM Wardlaw, IJ Deary

## Abstract

Cerebral small vessel disease (SVD) is a leading cause of vascular cognitive impairment, however the precise nature of SVD-related cognitive deficits, and their associations with structural brain changes, remain unclear. We combined computational volumes and visually-rated MRI markers of SVD to quantify total SVD burden, using data from the Lothian Birth Cohort 1936 (n=540; age:72.6±0.7 years). We found negative associations between total SVD burden and general cognitive ability (standardised β: −0.363; 95%CI: [−0.49, −0.23]; p(FDR)<0.001), processing speed (−0.371 [−0.50, −0.24]; p(FDR)<0.001), verbal memory (−0.265; [−0.42, −0.11]; p(FDR)=0.002), and visuospatial ability (−0.170; [−0.32, −0.02]; p(FDR)=0.029). Only the association between SVD burden and processing speed remained after accounting for covariance with general cognitive ability (−0.325; [−0.61, −0.04]; p(FDR)=0.029). This suggests that SVD’s association with poorer processing speed is not driven by, but is *independent* of its association with poorer general cognitive ability. Tests of processing speed may be particularly sensitive to the cognitive impact of SVD, but all major cognitive domains should be tested to determine the full range of SVD-related cognitive characteristics.

## 1. Introduction

Cerebral small vessel disease (SVD) is a leading cause of vascular cognitive impairment, contributing to multiple neurological disorders ranging from stroke, to mild cognitive impairment and dementia. Whereas current neuroimaging methods lack the spatial resolution to visualise the brain’s small vessels themselves, the downstream impact of their dysfunction is visible on brain imaging as white matter hyperintensities (WMH) and lacunes of presumed vascular origin, visible perivascular spaces (PVS) and cerebral microbleeds (Wardlaw et al., 2019). The presence and progression of radiological markers of SVD are frequently used as outcome measures in trials of interventions and treatments for SVD, however, the nature and extent of their associations with domain-specific cognitive outcomes remains unclear.

The majority of the literature examining SVD-related brain changes and cognitive ability focuses on individual radiological markers of SVD (most commonly WMH volume), thus failing to account for their potentially cumulative impact on cognitive performance. There is evidential support for considering the ‘total’ burden of SVD as a variable: different types of SVD lesions commonly occur together, are aetiologically related (Wardlaw et al., 2013), and associate with one another across a range of patient and healthy ageing populations (Ghaznawi et al., 2019, Potter et al., 2015). In recent years, studies have quantified the total burden of SVD with a 0-4 score, which allocates one point for the presence of WMH, PVS, lacunes or microbleeds (Al Olama et al., 2020, Banerjee et al., 2018, Del Brutto et al., 2018, Huijts et al., 2013, Uiterwijk et al., 2016). Whereas the 0-4 score can be calculated quickly from visual inspection of a brain scan, it lacks sensitivity to the range or severity of the individual SVD markers. To increase the fidelity of SVD burden quantification, two recent studies have developed continuous total SVD scores (Jokinen et al., 2020, Staals et al., 2015), finding negative associations between the continuous SVD burden score and domain-specific cognitive abilities. These studies also found that the magnitudes these associations surpassed those of models using a simple 0-4 SVD burden score (Staals et al., 2015) or individual MRI markers of SVD (Jokinen et al., 2020) as predictors of cognitive performance. However, research on cognitive ageing has established that age-related cognitive decline is not only observed across different domains of cognitive ability, but is *shared* between them owing to an overall decline in general cognitive ability (Salthouse, 2010, Tucker-Drob et al., 2019). A key outstanding question, therefore, is whether poor cognitive performance in certain cognitive domains is associated with general cognitive decline, or whether SVD has additional independent associations with specific domains of cognitive ability.

In this study we extend the work of Staals and colleagues (2015) by constructing a variable representing the total MRI-visible burden of SVD. We use structural equation modelling to combine continuous computational measures of white matter hyperintensities (WMH) and visible perivascular spaces (PVS), as opposed to previous studies that have employed ordinal measures derived from visual rating scales. We test whether the inclusion of these continuous SVD markers increases the sensitivity of the SVD score in its associations with general and domain-specific cognitive abilities. We then test whether poor performance in certain cognitive domains is associated with general cognitive decline, or whether SVD has additional independent associations with specific domains of cognitive ability. By gaining insight into the nature of the associations between the total brain burden of SVD and cognitive abilities, we aim to better characterise SVD-related cognitive impairment and facilitate its accurate measurement in trials and in clinical management.

## 2. Materials and Methods

### 2.1 Study cohort

Participants were members of the LBC1936, which has been described previously (Taylor et al., 2018). Briefly, the LBC1936 is a longitudinal follow-up to the Scottish Mental Survey of 1947, which assessed the cognitive ability of 70,805 11 year-old children, who were born in 1936 and were attending school in Scotland (Scottish Council for Research in Education, 1933). The present study includes participants from Wave 2 of the study, the first wave at which neuroimaging was carried out (usable neuroimaging data were available for n=680). Visible PVS are extremely small on neuroimaging (≤3 mm), therefore their computational detection is highly sensitive to noise and motion artefacts. Because of this, quality requirements for the MR images used in this study were high and images for 140 participants with available neuroimaging data could not be processed through the PVS pipeline (Ballerini et al., 2020). Reasons for this were failed registration of the centrum semiovale, noise or motion artefacts (which can have a similar appearance to PVS), or where small WMH were misclassified as PVS (Ballerini et al., 2020). Lack of computational PVS segmentation was the only factor that prevented inclusion in the study, thus the remaining 540 participants constitute our final sample. Approval for the LBC1936 was obtained from the Scotland A Research Ethics Committee for Scotland (07/MRE00/58). All participants gave written, informed consent.

### 2.2 MRI acquisition and radiological markers of SVD

Details of the MRI acquisition protocol have been published previously (Wardlaw et al., 2011). Briefly, participants were scanned using a GE Signa Horizon HDx 1.5 Tesla clinical scanner (General Electric, Milwaukee, WI) operating in ‘research mode’, equipped with a self-shielding gradient set (33 mT/m maximum gradient strength) and manufacturer supplied eight-channel phased-array head coil. Sequences acquired were T1-weighted (T1W), T2-weighted (T2W), T2*-weighted (T2*W) and fluid attenuated inversion recovery-weighted (FLAIR) images. MRI markers of SVD (WMH, PVS, lacunes and microbleeds) were measured using a combination of computational and visual rating methods, all performed blind to clinical and cognitive data (see Table 1; Wardlaw et al., 2011, 2013). In all analyses we divide WMH volume by total intracranial volume (TIV) to account for differences in head size.

**Table 1:** Definitions of key imaging features of SVD on structural MRI

| SVD feature | Definition and acquisition |
| --- | --- |
| White matter hyperintensities (WMH) | **Visual rating:** Periventricular and deep WMH were rated on the Fazekas scale (range 0-3) using FLAIR- and T2-weighted sequences (Fazekas et al., 1987).<br>**Computational volume:** WMH and total intracranial volumes (TIV) were measured using MCMxxVI (Valdés Hernández et al., 2010), a validated multispectral image processing method that combines T2, T2*W and FLAIR sequences for segmentation. All slices of all scans were checked by a trained observer and manually corrected, if necessary, to ensure that no true WMH had been omitted and to avoid including erroneous tissues in the WMH. |
| Visible perivascular spaces (PVS) | **Visual rating:** PVS in the basal ganglia were quantified using a previously described visual rating scale (Doubal et al., 2010) and were defined as small punctate or linear hyperintensities, in axial and longitudinal section respectively, on T2W that are <3mm in diameter. Larger PVS may be visible on T1W as decreased signal, but not visible on T2W or FLAIR (Wardlaw et al., 2011, 2013).<br>**Computational count:** PVS were computationally segmented in the native T2W space in the centrum semiovale using a recently validated technique (Ballerini et al., 2018, 2020) and are presented as total number of PVS. After segmentation of the PVS, all binary PVS masks (superimposed on T2-weighted images) were visually checked for the accurate quantification of PVS by a trained operator and were accepted or rejected blind to all other data. Where ambiguity arose, FLAIR and T1-weighted sequences were also checked (Ballerini et al., 2020). |
| Lacunes | Lacunes were classified as being present or absent and were defined as small (>3mm and <2cm in diameter) subcortical lesions of CSF-equivalent signal on T2W and decreased signal on T1W and FLAIR images in the white matter, basal ganglia, and brainstem (Wardlaw et al., 2011, 2013). |
| Cerebral microbleeds | Cerebral microbleeds were classified as being present or absent and were defined as small (<5mm), homogenous, round foci of low signal intensity on T2*W images in the white matter, basal ganglia, brain stem, cerebellum, and cortico-subcortical junction (Wardlaw et al., 2011, 2013). |

*Note:* Visual ratings were made by an experienced, registered neuroradiologist and a random 20% sample and any uncertain cases were independently checked by a second neuroradiologist, with disagreements resolved by consensus.

### 2.3 Cognitive data

Participants completed the Moray House Test No.12 (MHT), a test of general intelligence, at the age of 11 as part of the Scottish Mental Survey of 1947 (Scottish Council for Research in Education, 1933). In later life, participants completed a comprehensive battery of cognitive tests as part of the LBC1936, which we have grouped into three domains (processing speed, verbal memory, and visuospatial ability) according to prior work characterising their correlational structure (Tucker-Drob et al., 2014). The domain of processing speed includes Digit Symbol Substitution and Symbol Search from the Wechsler Adult Intelligence Scale-III (WAIS-III^UK^; Wechsler, 1998a) and two experimental tasks: Four Choice Reaction Time (Deary et al., 2001) and Inspection Time (Deary et al., 2004). Four Choice Reaction Time scores were multiplied by −1 so that higher scores indicated better performance. The domain of verbal memory includes Verbal Paired Associates (total score) and Logical Memory from the Wechsler Memory Scale III^UK^ (WMS-III^UK^; Wechsler, 1998b), and Digit Span (WAIS-III^UK^). Visuospatial ability includes Block Design, Matrix Reasoning (both WAIS-III^UK^) and Spatial Span (average of forwards and backwards; WMS-III^UK^). Scores on the ten cognitive tests included these domains were considered together as an indicator of general cognitive ability, given the well-replicated shared covariances of test scores across domains (Deary et al., 2010).

### 2.4 Covariates

Cognitive and neuroimaging data were acquired on two separate occasions. To account for variation in time intervals between these two occasions across the cohort, we adjusted the manifest cognitive variables for the difference in days between imaging and cognitive data acquisition outside of the SEM models (residualised using linear regression). We included age in years at the time of MRI, sex, vascular risk, the depression sub-score from the Hospital Anxiety and Depression Scale (HADS-D), and MHT score at age 11 (subsequently referred to as age-11 IQ) as covariates in all of our models. Vascular risk variables included self-reported history of hypertension (yes/no), diabetes mellitus (yes/no) and smoking status (ever/never); blood-derived glycated haemoglobin (% total HbA1c); blood-derived total cholesterol (mmol/l); and systolic and diastolic blood pressure (average of six readings: three seated and three standing), which were measured by trained nurses. We used confirmatory factor analysis (CFA) to construct a latent variable representing vascular risk as previously modelled in this cohort (Wardlaw et al., 2014) and extracted its factor score for inclusion as a covariate.

### 2.5 Statistical analysis

#### 2.5.1 Measurement models

We used CFA to construct a computationally-derived latent variable representing the total MRI-visible burden of SVD. This CFA assumed that the covariance among its indicators (WMH volume/TIV, PVS, lacunes, and microbleeds) was due to a single underlying factor (SVD), which is separate from unique and error variance in the four contributing variables. WMH volume/TIV and centrum semiovale PVS count were continuous computationally-derived variables, and lacunes and microbleeds were binary variables (i.e. present/absent), derived from visual assessment.

We also used CFA to reconstruct the latent total SVD variable based on ordinal visual scores used by Staals et al. (2015), which included deep and periventricular Fazekas scores as measures of WMH (both range 0-3), a visual rating scale for the assessment of PVS in the basal ganglia (range 0-4; Doubal et al., 2010), and counts of cerebral microbleeds and lacunes, which we converted to binary variables (present/absent) due to the low frequency of values greater than one.

#### 2.5.2 Multivariable SEM models

First, we specified separate linear regressions between the computationally-derived SVD burden variable (independent variable) and latent variables of general cognitive ability, processing speed, verbal memory, and visuospatial ability. These models tested the association between total SVD burden and the cognitive factors as described in the measurement models above. We included age, sex, vascular risk (extracted factor score), HADS-D score, and age-11 IQ as covariates in a step-wise manner to assess their impact on any associations. Covariates were free to correlate with one another and with total SVD burden, and were also regressed on the cognitive factor. To assess the extent to which these associations might be driven by the contribution of WMH to the SVD burden score, we also ran these models with WMH/TIV as the predictor and compared the effect size magnitudes from both sets of models using the Williams test (Williams, 1959), implemented in the *cocor* package (Diedenhofen and Musch, 2015) in R version 4.0.1 (R Core Team, 2020).

Next, we ran these analyses using the reconstructed total SVD variable as created by Staals et al. (2015) as the predictor. We compared the effect size magnitudes of these models with those of models including the computationally-derived SVD variable as a predictor, again using the Williams test.

General cognitive ability accounts for approximately 40% of the variability in performance on diverse batteries of cognitive tests (Carroll, 1993). Thus, given that cognitive domains are all substantially and positively correlated, in order to generate a domain-specific cognitive score, one must account for its covariance with other cognitive domains (i.e. general cognitive ability). To account for the confounding effects of general cognitive ability, we next tested associations between the computationally-derived SVD burden variable and a bifactor model of general cognitive ability (Fig. 1), which partitions variance in the cognitive test scores into that which contributes to general cognitive ability, and that which uniquely contributes to the domain-specific factors. The results of this model will indicate whether total SVD burden associates with any of the domain-specific cognitive scores *independently* of the variance that the domain-specific scores share (i.e. general cognitive ability). In this model we included age, sex, vascular risk HADS-D score, and age-11 IQ simultaneously as covariates.

**Figure 1:**
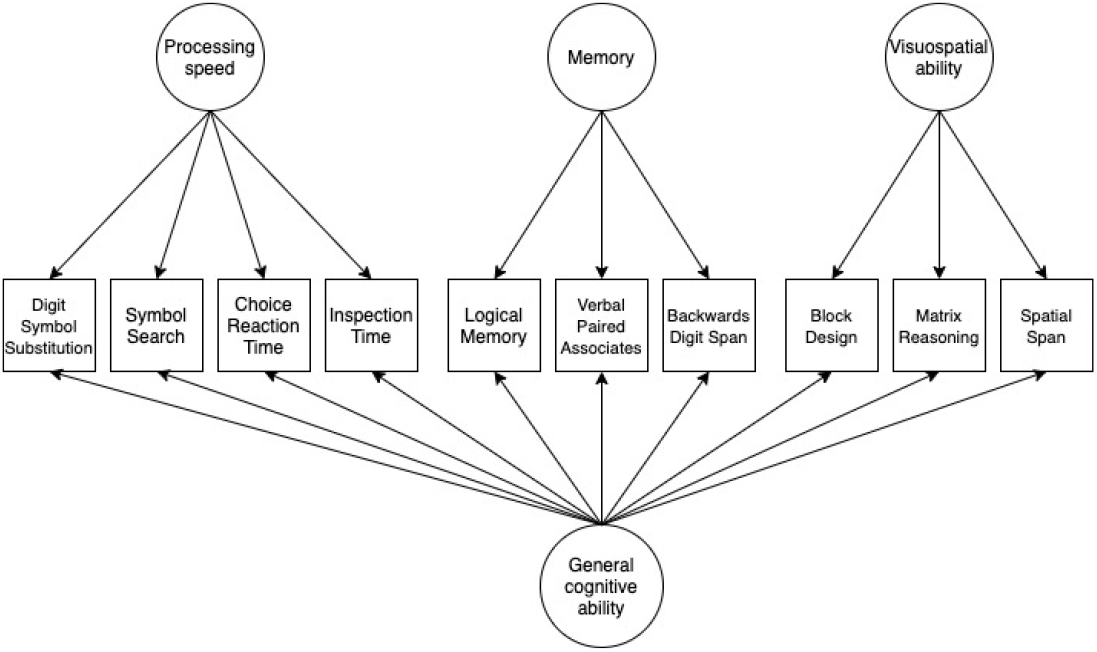
Diagram illustrating a bifactor model of general cognitive ability. *Figure 1 note:* Following conventional SEM notation, variables in squares were observed and measured, and variables in circles represent unmeasured latent variables. In this diagram, arrows indicate relationships between the underlying latent variables and the observed manifest variables.

The estimator for all multivariable SEM analyses was weighted least square mean and variance adjusted (WLSMV), which does not make distributional assumptions about observed variables. WLSMV uses logit link for continuous, and probit link for categorical variables. Model fit was assessed using four absolute fit indices: Root Mean Square Error of Approximation (RMSEA; <0.06 considered acceptable), Comparative Fit Index (CFI; >0.95 acceptable), Tucker-Lewis Index (TLI; >0.95 acceptable), and Standardized Root Mean Square Residual (SRMR; <0.08 acceptable). Pairwise present data were analysed due to the small amount of missing data (all cognitive variables had n ≥530). Data were analysed in MPlus version 8.3 (Muthén and Muthén, 1998-2017). We corrected *p*-values for multiple comparisons using the False Discovery Rate adjustment (FDR; Benjamini and Hochberg, 1995) with *p.adjust* in R version 4.0.1 (R Core Team, 2020).

### 2.6 Data availability

Data supporting this study are available upon reasonable request from the corresponding authors.

## 3. Results

### 3.1 Participant characteristics and SVD burden quantification

Characteristics of the study participants are presented in Table 2. The four MRI markers of SVD loaded significantly onto the computationally-derived SVD burden variable (Fig. 2) and the model fitted well (RMSEA=0.00; CFI=1.00; TLI=1.02; SRMR= 0.019). The four marker variables had moderate to large loadings on the latent SVD variable, which accounted for 25% of the variance in WMH/TIV, 10% of the variance in PVS, 32% of the variance in lacunes, and 18% of the variance in microbleeds.

**Table 2:** Characteristics of the study sample *Table 2 note*. CVD: Cardiovascular disease; HADS-D: depression sub-score of the Hospital Anxiety and Depression Scale; PVS visible perivascular spaces; TIV: total intercranial volume; WMH: white matter hyperintensities of presumed vascular origin.

|  | n | Mean (SD) unless otherwise stated |
| --- | --- | --- |
| **Sociodemographic** |  |  |
| Age, years | 540 | 72.6 (0.7) |
| Female, n (%) | 540 | 252 (46.7%) |
| Education, years | 540 | 10.9 (1.2) |
| **Vascular risk** |  |  |
| Hypertension history (yes), n (%) | 540 | 259 (48.0%) |
| Systolic blood pressure mmHg | 534 | 146.5 (18.0) |
| Diastolic blood pressure mmHg | 534 | 79.7 (18.0) |
| Diabetes history (yes), n (%) | 540 | 54 (10.0%) |
| HbA1c % total | 518 | 5.7 (0.6) |
| Cholesterol, mmol/l | 521 | 5.2 (1.1) |
| CVD history, n (%) | 540 | 154 (28.5%) |
| Smoking status, n (%) | 540 | Ever = 274 (50.7%)<br>Never = 266 (49.3%) |
| **Neurological/psychiatric** |  |  |
| Self-reported dementia, n (%) | 540 | 0 (0%) |
| Self-reported stroke, n (%) | 534 | 37 (10.5%) |
| HADS-D score | 540 | 2.5 (2.1) |
| **Neuroimaging** |  |  |
| Total WMH volume/TIV | 536 | 0.008 (0.009) |
| PVS count | 540 | 258.7 (94.6) |
| Lacunes, n (%) | 540 | Present = 28 (5.1%)<br>Absent = 512 (94.9%) |
| Microbleeds, n (%) | 540 | Present = 65 (12.0%)<br>Absent = 475 (88.0%) |
| **Cognitive** |  |  |
| Moray House Test score age-11 (max. 76) | 511 | 50.2 (11.9) |

**Figure 2:**
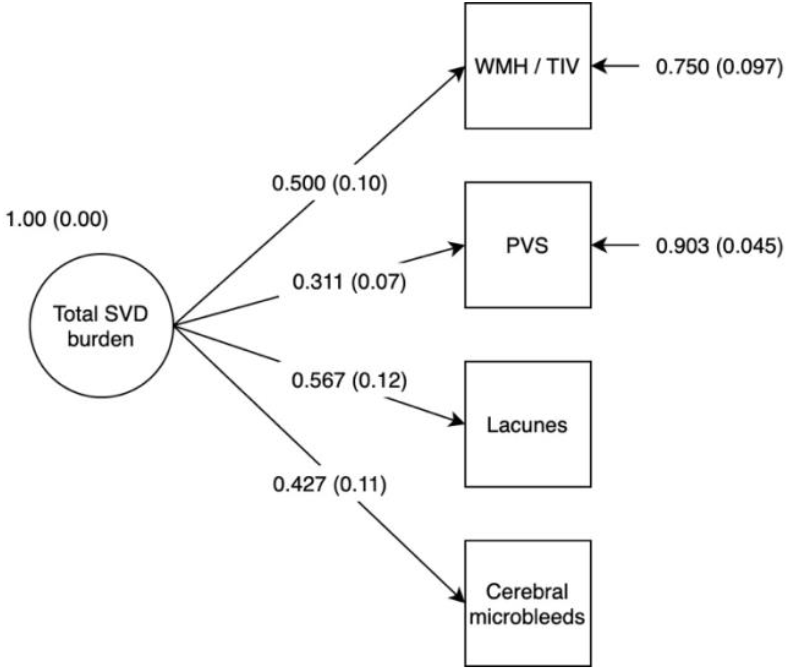
CFA diagram of latent variable representing total MRI-visible SVD burden. *Figure 2 note:* Latent variable representing total MRI-visible SVD burden. Estimator: WLSMV; RMSEA=0.00; CFI=1.00; TLI=1.02; SRMR=0.019. All indicators loaded significantly onto the factor at p<0.001. Standard errors are given in parentheses. For continuous variables (WMH and PVS) factor loadings represent standardised linear regression coefficients. For the binary variables (lacunes and microbleeds) factor loadings represent standardised probit regression coefficients. Factor loadings were freed for their interpretation. Following conventional SEM notation, variables in squares were observed and measured, and variables in circles represent unmeasured latent variables. Single headed arrows represent a relationship between two variables – in this model, this is either a linear or probit regression, with the arrow pointing towards the dependant variable.

### 3.2 Computationally-derived SVD burden score associates negatively with all cognitive domains

Total SVD burden demonstrated negative associations with all cognitive domains (Table 3). These associations remained significant after the inclusion of age, sex, vascular risk HADS-D score and age-11 IQ as covariates; covariate-adjusted absolute effect sizes range from −0.17 to −0.37. The latent variable representing total SVD burden accounted for 13% of the variance in general cognitive ability, 14% of the variance in processing speed, 7% of the variance in verbal memory, and 3% of the variance in visuospatial ability. Williams tests indicated that the magnitudes of these models (including all covariates) were significantly greater than those specifying WMH/TIV as the predictor: general cognitive ability (Williams’ one-sided *t*-value=−7.38; *p*<0.001), processing speed (*t*=−7.28; *p*<0.001); verbal memory (*t*=−5.06; *p*<0.001), visuospatial ability (*t*=−2.81; *p*=0.0051). Results of associations between WMH/TIV and cognitive factors are presented in Table S3.

**Table 3:**
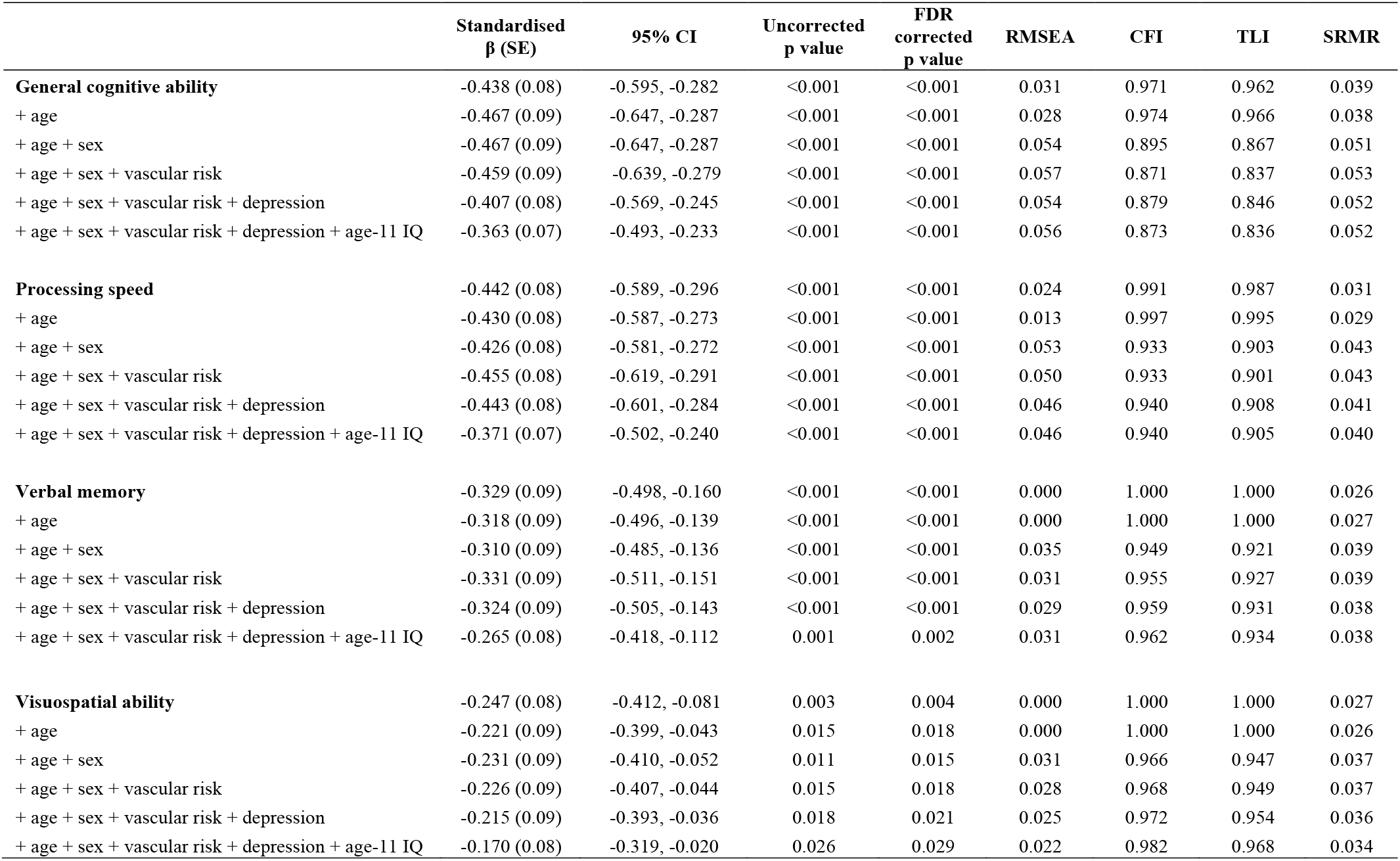
Associations between the computationally-derived total SVD burden variable and cognitive domains *Table 3 note*. N=540 for all analyses. CFI: Comparative Fit Index; RMSEA: Root Mean Square Error of Approximation; SRMR: Standardized Root Mean Square Residual; TLI: Tucker Lewis Index. After the inclusion of sex as a covariate in the models, the TLI and/or CFI fell below conventional thresholds (both >0.95). Off-diagonal values of the residual correlation matrix indicated that there were correlations between sex and the residuals of several manifest cognitive variables, which were unaccounted for in our model. When we specified regressions between sex and these residuals, the TLI and CFI reached acceptable levels. Combined with the good fit of our initial measurement models, this indicates that the lower CFI and TLI values of these models are due to unspecified correlations between sex and cognitive variables and are not due to model mis-specification.

Similarly, the reconstructed total SVD burden variable used by Staals et al. (2015) associated negatively with all cognitive domains (Table 4). Williams tests indicated that the magnitudes of these models (including all covariates) were significantly smaller than those using the computationally-derived SVD burden variable as the predictor: general cognitive ability (Williams’ one-sided *t*-value=−6.25; *p*<0.001), processing speed (*t*=−6.11; *p*<0.001); verbal memory (*t*=−4.48; *p*<0.001), visuospatial ability (*t*=−2.51; *p*=0.0123).

**Table 4:**
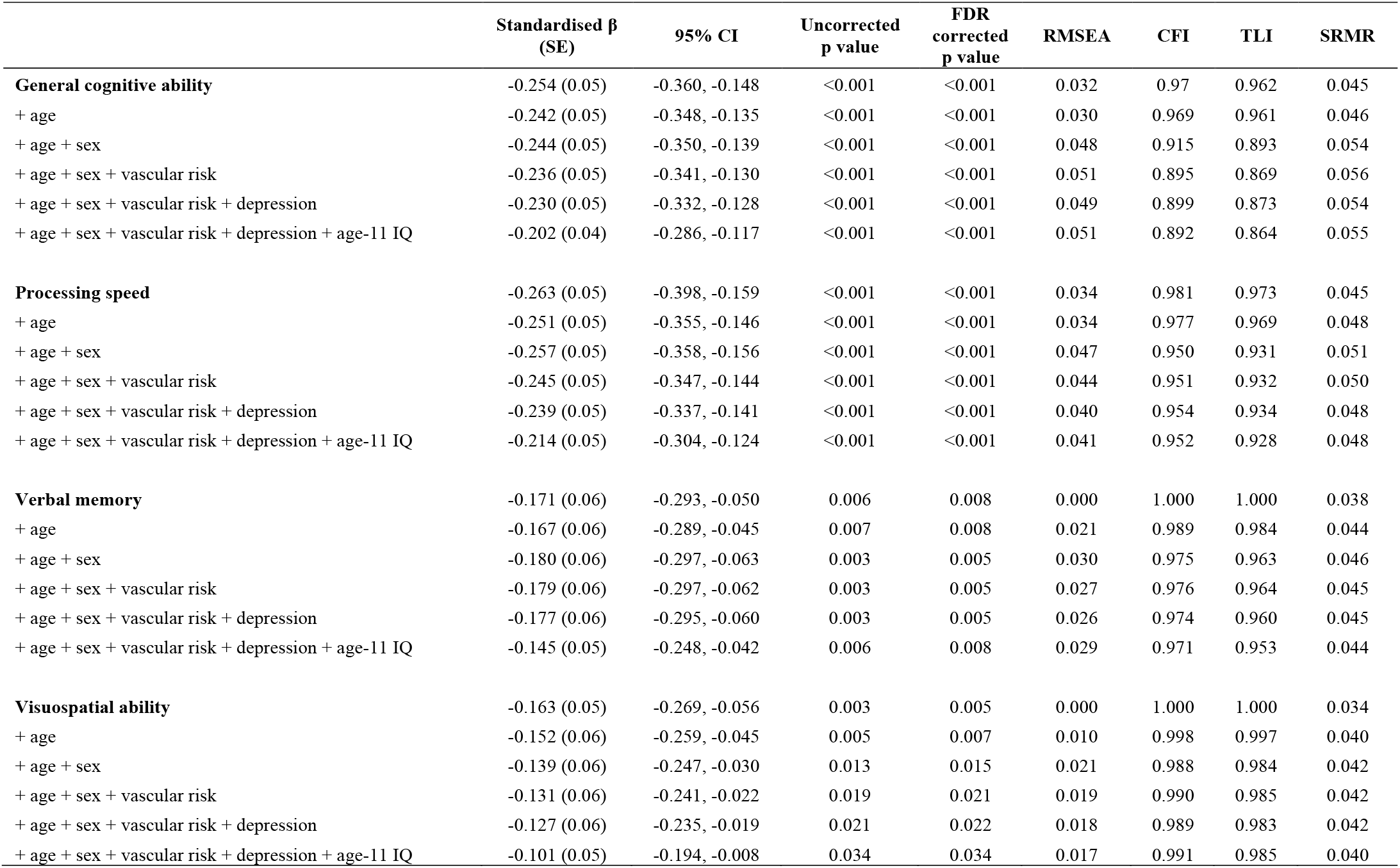
Associations between the reconstructed SVD burden variable used by Staals et al. and cognitive factors *Table 4 note*. N=540 for all analyses. CFI: Comparative Fit Index; RMSEA: Root Mean Square Approximation; SRMR: Standardized Root Mean Square Residual; TLI: Tucker Lewis Index.

### 3.3 Computationally-derived SVD burden score shows a specific and independent association with processing speed

The multivariable bifactor model, performed to test associations between the computationally-derived SVD burden score and domain-specific cognitive abilities independently of general cognitive ability, fitted well (Fig. 3; RMSEA=0.030; CFI=0.970; TLI= 0.952; SRMR=0.044). Total SVD burden was negatively associated with general cognitive ability (standardised β: −0.224; 95%CI: [−0.40, −0.05]; *p(FDR)*=0.016) and processing speed (−0.325; [−0.61, −0.04], *p(FDR)*=0.029), and accounted for 5% of the variance in general cognitive ability and 11% of the variance in residual processing speed. There were no significant associations between total SVD burden and verbal memory (−0.133, [−0.32, 0.05], *p(FDR)*=0.166) or visuospatial ability (0.075; [−0.25, 0.40], *p(FDR)*=0.654). The latter result may be due to the heavy loading of the visuospatial tests onto general cognitive ability, which left relatively little variance for the independent visuospatial factor.

**Figure 3:**
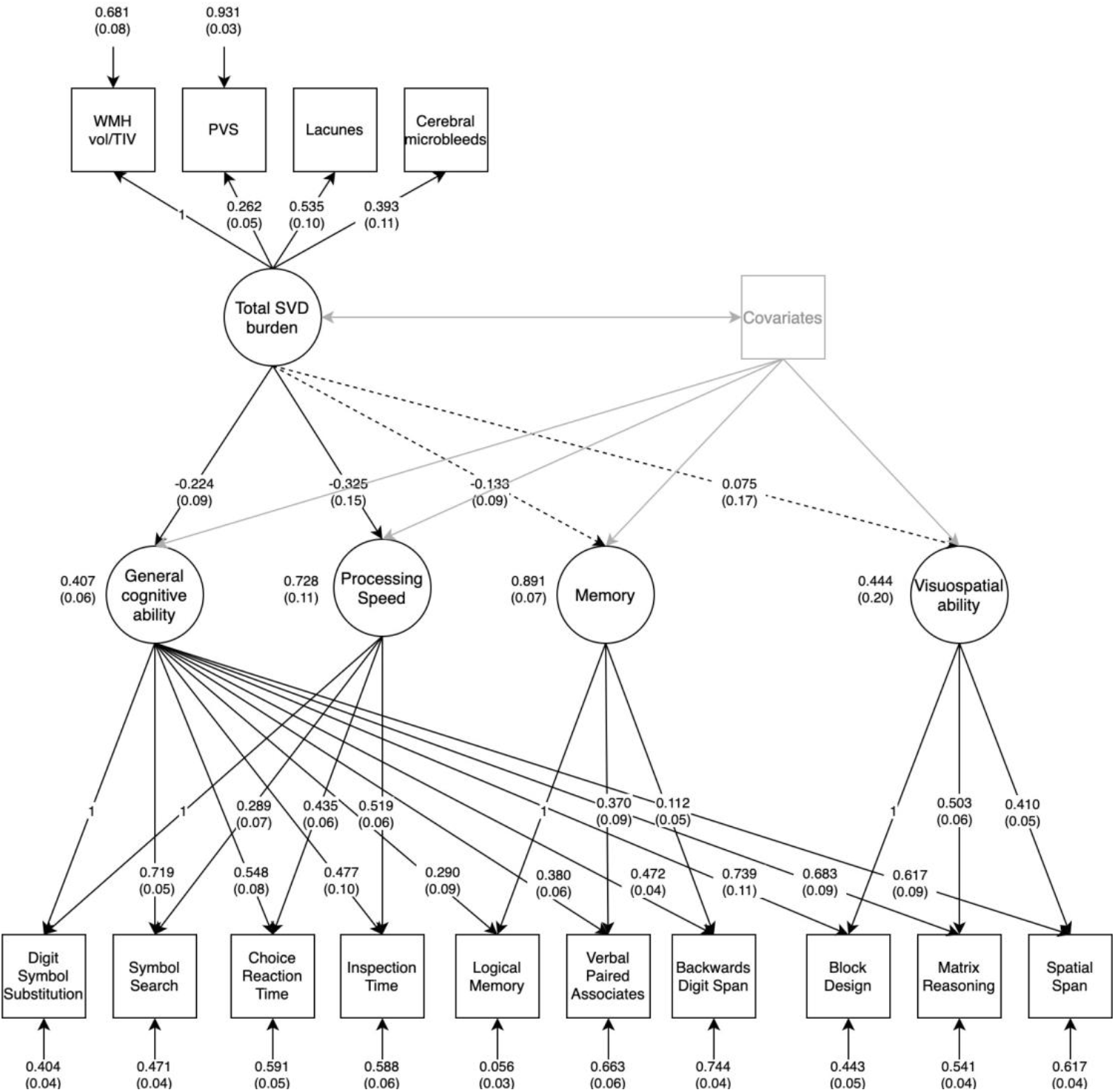
SEM diagram illustrating associations between SVD burden and a bifactor model of general cognitive ability. *Figure 3 note:* Associations between total SVD burden and a bifactor model of cognitive ability (n=540). Estimator: WLSMV. Fit indices: RMSEA=0.030; CFI=0.970; TLI=0.952; SRMR=0.044. Solid black lines between total SVD burden and cognitive factors represent significant associations and dashed lines represent non-significant associations after FDR correction. Factor loadings of these four associations are standardised linear regression coefficients and standard errors are shown in parentheses. Age at time of MRI, sex, vascular risk, HADS depression sub-score, and age-11 IQ were included as covariates and were free to correlate with one another

## 4. Discussion

To date, the majority of studies have used individual MRI markers of SVD, or a simple 0-4 sum score to quantify SVD burden. In this study of 540 community-dwelling older adults, we combined computational and visually-rated MRI markers of SVD (including for the first time, a continuous measure of PVS) to estimate a continuous latent variable representing total MRI-visible SVD burden. In doing so, we were able to increase the fidelity with which SVD burden is quantified, relative to a previous method which relied on visually-rated SVD markers only. The results of our analyses indicated that a higher SVD burden was not only associated with poorer general cognitive ability and processing speed, as current consensus statements suggest (Peng et al., 2019, Rosenberg et al., 2016), but also with poorer memory and visuospatial ability. We then accounted for the covariance between cognitive domain scores and general cognitive ability in a bifactor model, finding that total SVD burden was associated with processing speed not only due to, but *in addition* to its association with poorer general cognitive ability. A comparison of the covariate-adjusted effect sizes for associations between total SVD burden and processing speed before accounting for general cognitive ability (simple regression model standardised β: −0.371) and after accounting for general cognitive ability (bifactor model standardised β: −0.325), suggests that approximately 12% of the variance in processing speed is accounted for by general ability. Therefore, failing to account for covariance with general cognitive ability could lead to an overestimation of effect sizes between SVD burden and processing speed.

Slowed processing speed and poor executive function are often considered to be the hallmark cognitive features of SVD, with little attention given to other cognitive domains. However, our results suggest that alongside slowed processing speed, SVD burden is also related to poorer performance on tests of memory and visuospatial ability, even in a cohort of individuals with mild, non-clinical presentations of SVD. We found associations between SVD burden and verbal memory and visuospatial ability in separate regression models, but not in the bifactor model. This suggests that negative associations between SVD and both memory and visuospatial ability may be a consequence of the negative association between SVD and general cognitive ability. It has been suggested that poor performance on tests of memory and visuospatial ability (two cognitive abilities subtended by specific cortical areas) could result from the disruption of white matter connections between cortical and subcortical regions (Tuladhar et al., 2015). Our data support this notion and suggest that damage to these connections may be part of a more general, diffuse process.

Slowed information processing speed is recognised as a key feature of SVD. However, processing speed test scores are a chimera; that is, part of the variation in processing speed is due to its association with general cognitive ability. Therefore, it was previously unknown whether the apparent association between SVD and slowed processing speed was due to SVD’s impact on general cognitive ability, or whether SVD may have specific and independent effects on processing speed. Here, we have removed the general cognitive ability variance from processing speed (and other cognitive domains), which affords a better test of any SVD-processing speed association. Our results suggested that the association between SVD burden and processing speed was *independent* of general cognitive ability, thus favouring the latter hypothesis. To the best of our knowledge, this is the first study to demonstrate an association between SVD burden and processing speed independently of the shared variance between cognitive test scores, which acts as a confound. Typically, this confound remains unaccounted for, thus previous studies that have reported associations between SVD and scores from tests of processing speed could be misleading. Processing speed is often regarded as having a special status among the domains of cognitive ability; it is typically the first domain affected by age, and as performance on tasks in a variety of cognitive domains relies on information processing, its early decline may lead change in other domains (Finkel et al., 2007). It follows that networks supporting processing speed appear to be distributed throughout the brain: previously in the LBC1936, poorer processing speed was associated with age-related reductions in white matter microstructural integrity across the whole brain, and in broad regions of interest (Deary et al., 2006, Kuznetsova et al., 2016, Penke et al., 2010). That total SVD burden may have a specific impact on processing speed, independent of its effect on general cognitive ability, further suggests that SVD-related brain changes are widespread, rather than tract-specific.

Previously, in 680 participants from the LBC1936 Staals et al. (2015) found no associations between their total SVD burden score (derived from visual rating scales of individual MRI markers of SVD) and a composite score of processing speed. This composite score of processing speed was extracted from a bifactor model of general cognitive ability, so as in our study, was independent of general cognitive ability. There are several key differences between the present study and that of Staals et al. (2015), which may account for our differing results. First, we were unable to include 140 of Staals’ 680 participants due to MRI noise or motion artefacts, which precluded the quantification of the computational PVS measure. As a higher burden of imaging artefacts likely reflect poorer brain health, the range of SVD severity is potentially reduced in our sample. The anticipated effect of this would be a reduction in the magnitude of observed effect sizes between SVD burden and cognitive outcomes, however this was not the case, so our smaller, less noisy sample is unlikely to be responsible for our differing results. Second, we used continuous as opposed to ordinal MRI data for two key SVD features (WMH and PVS) of our total SVD burden variable. In separate regression models, the reconstructed SVD variable used by Staals et al. was negatively associated with all cognitive domains that we tested, but the effect size magnitudes of these models were significantly smaller than those using the computationally-derived SVD score as a predictor. This suggests that incorporating continuous measurements of WMH and PVS into the original total SVD burden score increased the fidelity of the SVD burden measure, revealing associations with cognitive outcomes which were previously unobserved. The computationally-derived SVD variable also demonstrated greater predictive power in its associations with cognitive outcomes than WMH/TIV, suggesting that some of these effects may be missed when using WMH volume as the sole predictor of cognitive performance.

Whereas tests of processing speed may be particularly sensitive to, and possibly an early indicator of, the cognitive impact of SVD (Deary et al., 2019), our findings suggest that SVD burden also associates with poorer performance on tests of verbal memory and visuospatial ability. Future research studies and clinical trials assessing domain-specific cognitive outcomes in SVD should also assess all major domains of cognitive ability in order to capture a more accurate picture of SVD-related cognitive impairments. We have also demonstrated the benefit of constructing a computationally-derived variable of total SVD burden, over one constructed using visually-rated MRI data alone. A latent variable of SVD burden using continuous MRI data may be useful in a research setting for testing associations between SVD burden and clinically-relevant outcomes, however, further interrogation of the latent SVD burden variable is required before it could be considered for use as a marker of SVD severity in a clinical trial setting. We are yet to examine, for example, how the latent SVD variable might change over time, or whether it can be constructed in clinical populations with more substantial burden of SVD or more complex multi-morbidities, such as Alzheimer’s disease. The consistency of latent SVD variables using computationally-derived data might also vary according to scanning parameters or methods used to quantify the radiological markers of SVD.

Strengths of this study include our large sample size, extensive cognitive testing, and detailed assessment of biomarker variables, which enabled us to account for a broad range of vascular risk factors. Additionally, as childhood IQ accounts for approximately 50% of the variance in cognitive ability in later life (Deary, 2014), a further strength of this study is our ability to account for this confound by including age-11 IQ as a covariate in our analyses. Age-11 IQ had the greatest impact of any of the covariates we included, attenuating the standardised betas of the associations between total SVD burden and later-life cognitive abilities by an average of about .04. Limitations of this study include our reliance on binary measurements of lacunes and microbleeds, which were used due to the scarcity of participants in our sample with more than one lacune or microbleed. In a sample of participants with more substantial SVD pathology, it might be desirable to model these variables as count data, as the reduction of highly variable data into binary outcomes results in the loss of information. Computational continuous measures of lacunes and microbleeds are also feasible, however, it is not yet clear whether these should be expressed as a total volume or count; microbleeds may be contaminated with other mineral deposits, and as the least frequent SVD lesions, their contribution to the total SVD burden is well captured in a binary score. A valid computational measure would likely further increase the sensitivity of the latent SVD variable and should be tested in future, especially in more diseased populations likely to have more lacunes and microbleeds. A further limitation of this study is that the computationally-derived SVD variable incorporated a measure of PVS in the centrum semiovale, whereas the reconstructed SVD score used by Staals et al. (2015) incorporated a measure of PVS in the basal ganglia. Whereas PVS in the centrum semiovale are related to cerebral amyloid angiopathy (CAA) pathology, they are also present in sporadic SVD; in the LBC1936, visual ratings of PVS in the centrum semiovale and basal ganglia correlate with one another (*r*=0.40; *p*<0.001) and strong associations between computational PVS count and other markers of SVD, such as Fazekas scores and WMH volumes, have previously been reported (Ballerini et al., 2020). However, the associations that we observed between SVD burden and cognitive ability in the domains of processing speed, verbal memory and visuospatial ability in the LBC1936 support the suggestion that SVD affects multiple cognitive domains before clinical presentation.

We constructed a computationally-derived variable representing the total MRI-visible burden of SVD using continuous scores of WMH and PVS and binary ratings of lacunes and microbleeds. SVD burden associated negatively with verbal memory and visuospatial ability, but this is likely due to SVD’s association with general cognitive ability. SVD burden was also negatively associated with processing speed, but this association was found to be independent of poorer general cognitive ability. Future research studies and clinical trials monitoring cognitive outcomes in SVD should assess the domains of memory and visuospatial ability in addition to processing speed, in order to capture a fuller and more clinically-relevant picture of SVD-related cognitive abilities.

## Data Availability

Data supporting this study are available upon reasonable request from the corresponding authors.

## Acknowledgements

The authors thank all participants of the LBC1936 who have contributed, and continue to contribute to the ongoing study. We thank the radiographers, nurses and Lothian Birth Cohort 1936 research team members who collected, entered and checked data used in this manuscript. We also thank Julie Staals and Tom Booth for their constructive comments on earlier versions of the manuscript. The LBC1936 is supported by Age UK [MR/M01311/1] (http://www.disconnectedmind.ed.ac.uk) and the Medical Research Council [G1001245/96099]. LBC1936 MRI brain imaging was supported by Medical Research Council (MRC) grants [G0701120], [G1001245], [MR/M013111/1] and [MR/R024065/1]. OKLH is funded by the University of Edinburgh College of Medicine and Veterinary Medicine as part of the Wellcome Trust 4-year PhD in Translational Neuroscience at the University of Edinburgh. SRC, JMW, IJD, SMM and LB were supported by MRC grants [MR/M013111/1] and [MR/R024065/1]. SRC and IJD are additionally supported by a National Institutes of Health (NIH) research grant R01AG054628, and IJD was also supported by the Dementias Platform UK [MR/L015382/1]. MVH was supported by the Row Fogo Charitable Trust [BROD.FID3668413]. JMW is supported by the European Union Horizon 2020, (PHC-03-15, project no 666881), ‘SVDs@Target’, the Fondation Leducq Transatlantic Network of Excellence for the Study of Perivascular Spaces in Small Vessel Disease (ref no. 16 CVD 05), and the UK Dementia Research Institute which receives its funding from DRI Ltd, funded by the UK Medical Research Council, Alzheimer’s Society and Alzheimer’s Research UK.

