## Supplementary Tables 1, 2 and 3 for "Associations between total MRI-visible small vessel disease burden and domain-specific cognitive abilities in a community-dwelling older-age cohort"

**Table S1:** Correlation matrix (Pearson coefficients) of MRI variables contributing to the computationally-derived total SVD burden variable (pg. 2)

**Table S2:** Correlation matrix (Pearson coefficients) of cognitive variables measured in older age (pg. 3)

**Table S3:** Associations between the total WMH volume/TIV and cognitive factors (pg. 4)

**Table S1**: Correlation matrix (Pearson coefficients) of MRI variables contributing to the computationally-derived total SVD burden variable

| | WMH/TIV | Computational PVS count | Lacunes (present/absent) | Microbleeds (present/absent) |
|---|---|---|---|---|
| WMH/TIV | - | | | |
| Computational PVS count | 0.16 | - | | |
| Lacunes (present/absent) | 0.21 | 0.06 | - | |
| Microbleeds (present/absent) | 0.16 | 0.10 | 0.09 | - |

**Table S2**: Correlation matrix (Pearson coefficients) of cognitive variables measured in older age

| | Symbol Search | Digit Symbol | Choice Reaction Time | Inspection Time | Verbal Paired Associates | Logical Memory | Backwards Digit Span | Matrix Reasoning | Block Design | Spatial Span |
|---|---|---|---|---|---|---|---|---|---|---|
| Symbol Search | - | | | | | | | | | |
| Digit Symbol | 0.64 | - | | | | | | | | |
| Choice Reaction Time | 0.49 | 0.52 | - | | | | | | | |
| Inspection Time | 0.35 | 0.42 | 0.40 | - | | | | | | |
| Verbal Paired Associates | 0.25 | 0.29 | 0.20 | 0.22 | - | | | | | |
| Logical Memory | 0.29 | 0.33 | 0.23 | 0.18 | 0.53 | - | | | | |
| Backwards Digit Span | 0.32 | 0.34 | 0.21 | 0.22 | 0.27 | 0.31 | - | | | |
| Matrix Reasoning | 0.37 | 0.35 | 0.22 | 0.28 | 0.34 | 0.34 | 0.32 | - | | |
| Block Design | 0.46 | 0.41 | 0.26 | 0.29 | 0.28 | 0.27 | 0.26 | 0.53 | - | |
| Spatial Span | 0.37 | 0.30 | 0.29 | 0.28 | 0.21 | 0.23 | 0.34 | 0.39 | 0.47 | - |

**Table S3:** Associations between the total WMH volume/TIV and cognitive factors

| | Standardised β (SE) | 95% CI | uncorrected p value | FDR corrected p value | $X^2$ | RMSEA | CFI | TLI | RMSEA |
|---|---|---|---|---|---|---|---|---|---|
| **General cognitive ability** | -0.272 (0.05) | -0.334, -0.154 | <0.001 | <0.001 | 0.000 | 0.06 | 0.953 | 0.933 | 0.038 |
| + age | -0.224 (0.05) | -0.315, -0.133 | <0.001 | <0.001 | 0.000 | 0.053 | 0.955 | 0.938 | 0.036 |
| + age + sex | -0.229 (0.05) | -0.320, -0.139 | <0.001 | <0.001 | 0.000 | 0.077 | 0.890 | 0.858 | 0.053 |
| + age + sex + vascular risk | -0.222 (0.05) | -0.313, -0.132 | <0.001 | <0.001 | 0.000 | 0.076 | 0.873 | 0.841 | 0.055 |
| + age + sex + vascular risk + depression | -0.220 (0.05) | -0.309, -0.131 | <0.001 | <0.001 | 0.000 | 0.071 | 0.875 | 0.848 | 0.053 |
| + age + sex + vascular risk + depression + age-11 IQ | -0.190 (0.04) | -0.265, -0.114 | <0.001 | <0.001 | 0.000 | 0.074 | 0.865 | 0.839 | 0.055 |
| **Processing speed** | -0.239 (0.05) | -0.328, -0.150 | <0.001 | <0.001 | 0.010 | 0.061 | 0.985 | 0.969 | 0.027 |
| + age | -0.221 (0.05) | -0.310, -0.131 | <0.001 | <0.001 | 0.057 | 0.041 | 0.989 | 0.981 | 0.022 |
| + age + sex | -0.232 (0.05) | -0.319, -0.144 | <0.001 | <0.001 | 0.000 | 0.074 | 0.944 | 0.923 | 0.044 |
| + age + sex + vascular risk | -0.225 (0.05) | -0.313, -0.137 | <0.001 | <0.001 | 0.000 | 0.087 | 0.899 | 0.869 | 0.053 |
| + age + sex + vascular risk + depression | -0.221 (0.04) | -0.307, -0.135 | <0.001 | <0.001 | 0.000 | 0.076 | 0.907 | 0.885 | 0.048 |
| + age + sex + vascular risk + depression + age-11 IQ | -0.201 (0.04) | -0.280, -0.123 | <0.001 | <0.001 | 0.000 | 0.078 | 0.901 | 0.882 | 0.048 |
| **Verbal memory** | -0.158 (0.05) | -0.257, -0.058 | 0.002 | 0.003 | 0.497 | 0.000 | 1.000 | 1.000 | 0.011 |
| + age | -0.152 (0.05) | -0.254, -0.051 | 0.003 | 0.004 | 0.211 | 0.029 | 0.992 | 0.983 | 0.017 |
| + age + sex | -0.176 (0.05) | -0.274, -0.077 | <0.001 | <0.001 | 0.011 | 0.052 | 0.954 | 0.931 | 0.032 |
| + age + sex + vascular risk | -0.175 (0.05) | -0.274, -0.076 | 0.001 | 0.002 | 0.000 | 0.088 | 0.823 | 0.758 | 0.052 |
| + age + sex + vascular risk + depression | -0.174 (0.05) | -0.272, -0.076 | 0.001 | 0.002 | 0.000 | 0.078 | 0.825 | 0.775 | 0.048 |
| + age + sex + vascular risk + depression + age-11 IQ | -0.141 (0.05) | -0.230, -0.052 | 0.002 | 0.003 | 0.000 | 0.086 | 0.835 | 0.796 | 0.049 |
| **Visuospatial ability** | -0.159 (0.05) | -0.256, -0.063 | 0.001 | 0.002 | 0.400 | 0.000 | 1.000 | 1.000 | 0.012 |
| + age | -0.142 (0.05) | -0.239, -0.045 | 0.004 | 0.005 | 0.599 | 0.000 | 1.000 | 1.000 | 0.012 |
| + age + sex | -0.126 (0.05) | -0.223, -0.029 | 0.011 | 0.013 | 0.342 | 0.015 | 0.997 | 0.996 | 0.025 |
| + age + sex + vascular risk | -0.121 (0.05) | -0.218, -0.024 | 0.015 | 0.016 | 0.000 | 0.076 | 0.902 | 0.866 | 0.049 |
| + age + sex + vascular risk + depression | -0.117 (0.05) | -0.213, -0.022 | 0.016 | 0.017 | 0.000 | 0.067 | 0.905 | 0.878 | 0.044 |
| + age + sex + vascular risk + depression + age-11 IQ | -0.099 (0.04) | -0.183, -0.014 | 0.022 | 0.022 | 0.000 | 0.065 | 0.922 | 0.904 | 0.044 |

*Note.* N=536 for all analyses. CFI: Comparative Fit Index; RMSEA: Root Mean Square Approximation; SRMR: Standardized Root Mean Square Residual; TIV: total intracranial index; TLI: Tucker Lewis Index; WMH: white matter hyperintensities. After the inclusion of sex as a covariate in the models for general cognitive ability, processing speed and verbal memory, the TLI and/or CFI fell below conventional thresholds (both >0.95). Off-diagonal values of the residual correlation matrix indicated that there were correlations between sex and the residuals of several manifest cognitive variables, which were unaccounted for in our model. When we specified regressions between sex and these residuals, the TLI and CFI reached acceptable levels. Combined with the good fit of our initial measurement models, this indicates that the lower CFI and TLI values of these models are due to unspecified correlations between sex and cognitive variables and are not due to model mis-specification. Model estimator was Maximum Likelihood, therefore $X^2$ is also included as a measure of model fit.
